# Acute and Longitudinal Effects of Sport-related Concussion on Reactive Balance

**DOI:** 10.1101/2024.03.28.24305029

**Authors:** Cecilia Monoli, Amanda Morris, Regan Crofts, Nora F. Fino, Tessa L. Petersell, Trevor Jameson, Leland E. Dibble, Peter C. Fino

## Abstract

Postural instability is a common observation after concussions, with balance assessments playing a crucial role in clinical evaluations. Widely used post-concussion balance tests focus primarily on static and dynamic balance, excluding the critical aspect of reactive balance. This study investigated the acute and longitudinal effects of concussion on reactive balance in collegiate athletes. The assessments were conducted at pre-season baseline and 4 post-concussion timepoints: acute, pre-return-to-play, post-return-to-play, and six months post-concussion. The instrumented-modified Push and Release test measured reactive balance. Longitudinal effects of concussions on time to stability and step latency metrics were investigated applying Generalized Estimating Equations. Acutely after concussion, athletes demonstrated impaired reactive balance, indicated by longer times to stability, in dual-task conditions (*p*= 0.004). These acute impairments were transient and recovered over time. Exploratory analyses revealed that athletes who sustained their first lifetime concussion exhibited both acute (*p* = 0.037) and longitudinal (*p* = 0.004 at post-return-to-play) impairments in single– and dual-task compared to controls with no lifetime concussion. This comprehensive evaluation provides insights into the multifaceted nature of post-concussion impairments and emphasizes the importance of considering cognitive demand and history of concussions in assessing athletes’ balance.

## INTRODUCTION

Postural instability is frequently observed after concussion^1^, highlighting the pivotal role of balance tests in clinical evaluations for this condition. Nevertheless, widely employed post-concussion balance assessments, such as the Balance Error Scoring System (BESS) and timed tandem gait (TTG), provide a limited perspective on balance^2^. A comprehensive understanding of balance includes several factors that influence individuals’ ability to stand, move, react, and engage with their surroundings^3,4^. These factors manifest within three primary domains of balance activities: static balance, dynamic balance, and reactive balance^5^. In concussion evaluations, the BESS test specifically targets static balance, defined as the ability to maintain equilibrium and control the center of mass in relation to a fixed base of support^6^. The TTG test assesses dynamic balance, described as the capacity to maintain equilibrium during motion or voluntary transitional movements^7^. Notably, the evaluation of reactive balance, which encompassing movements and postural responses to unpredictable disturbances for the restoration of equilibrium and stability^8^, is absent from post-concussion assessment. Given the importance of reactive balance for adapting to complex environments^9^ and its association with prospective musculoskeletal injuries in elite athletes^10^, there is a pressing need for further understanding the immediate and long-term impacts of concussions on reactive balance.

Reactive balance relies on interactions among spinal circuits, the brainstem, and the cerebral cortex^11^ to coordinate responses to sudden unexpected alterations in the base of support or center of mass kinematics. In contrast to static and dynamic balance tasks, reactive balance is initiated faster than voluntary movement and leaves little time for iterative corrections^5,12^. Instead, responses can be primed based on experiential learning in advance of the perturbation, and triggered (i.e., executed) promptly upon a loss of balance. While this priming involves cognitive resources^12^, measures of reactive balance are independent from computerized cognitive tests, clinical measures of reaction time, and measures of static and dynamic balance commonly used in concussion evaluations^13^. Consequently, evaluating reactive balance introduces a distinct dimension that remains independent of these conventional clinical metrics.

The limited prior work on reactive balance post-concussion has predominantly employed standing platform perturbations to investigate the cross-sectional and chronic effects of concussion (i.e., mild traumatic brain injury) on reactive balance performance^5^. Pan et al.^14^ provided preliminary evidence that individuals experiencing persistent symptoms after concussion exhibit impaired reactive balance in response to a sliding platform perturbation. However, these deficits were evident only among those with ongoing symptoms of disequilibrium at various chronicity, spanning from seven months to seven years; asymptomatic individuals with a history of concussion did not display reactive balance impairments. This heterogeneous time since concussion complicates our understanding of reactive balance post-concussion, including the acute and time-varying recovery of reactive balance.

Therefore, the purpose of this study was to assess the acute and longitudinal effects of concussion on reactive balance. Following a pre-registered protocol^15^, reactive balance of collegiate athletes with a recent concussion and matched controls was assessed at four clinically relevant timepoints: acutely after injury (<72 hours), when asymptomatic and cleared to begin the return-to-play (RTP) protocol (Pre-RTP), after completing the RTP protocol and cleared to return to full sport participation (Post-RTP), and six months after injury (6-Month). Additionally, a subset of athletes was tested upon arrival on campus before the season and any concussion event (Baseline), allowing for pre-to-post injury comparisons. The assessments were administered alongside the protocols of the Pac-12 CARE Affiliated Program^16^. The RTP protocol followed the guidelines set by the University of Utah Concussion Management Plan and the Sport Concussion Assessment Tool – 5 (SCAT5)^17^, a six-step process to monitor athlete’s concussion symptoms, cognitive and balance impairment.

We hypothesized that (H1a) athletes with acute concussion will take longer to regain balance during reactive balance tasks and (H1b) will display larger deficits in reactive balance during dual-tasks compared to healthy controls; (H2) athletes with a recent concussion will improve reactive balance throughout the assessments, but will still demonstrate persistent longitudinal deficits compared to the controls.

## MATERIALS and METHODS

### Participants

The study involved NCAA Division I student-athletes currently enrolled at the University of Utah, across the sanctioned sports, aged between 18 and 30. Exclusion criteria disqualified individuals that had a concussion within the past year prior to their enrollment, lower extremity surgeries within the previous two years, or planned surgeries that would affect their ability to participate in their sport. Additionally, individuals with a documented history of vestibular or somatosensory pathology were excluded. The testing procedures took place at the Athletic Training Clinics of the University of Utah in Salt Lake City, Utah. Prior to participation, all participants underwent written informed consent in accordance with a protocol approved by the Institutional Review Board.

All concussion subjects were diagnosed by a team physician and reported to the study team by the team’s athletic trainer(s). The team’s athletic trainer recommended control subjects among the concussion subject’s teammates, matched by sex, age, playing position, and skill level (in order of decreasing priority). Demographic data (age, sex, race, ethnicity, height, weight, and body mass index (BMI)), athletic activities (e.g., sport), lower extremity injury history, and self-reported lifetime concussion history were collected for each participant before completing any mobility and balance assessment.

Participants completed testing at five timepoints: at the start of intercollegiate competition at the University of Utah, typically as a freshmen or upon arriving on campus as a transfer (Baseline); within 72 hours after a concussion (Acute); within 72 hours of beginning the RTP protocol (Asymptomatic/Pre-RTP); within 72 hours of being cleared for unrestricted return to competition (Post-RTP); and 6 months after the initial injury (6-Month). Notably, not every participant was present at each of the scheduled assessment timepoints due to scheduling conflicts and referral delays.

Our enrollment goal was 40 participants per group at each timepoint ^15^. The sample size was calculated from differences between symptomatic concussed subjects and healthy controls in response to a tethered pull perturbation^14^. Using an estimated difference of 1.7 standard deviations (Cohen *d* = 1.7) between concussed subjects and healthy controls, we estimated 99% power to detect group differences at acute assessment with 40 subjects per group. Using an estimated effect size of Cohen *d* = 0.72 between asymptomatic (Pre-RTP timepoint) athletes and healthy controls, we estimated 74% power to detect group differences at the primary Post-RTP timepoint with 40 subjects per group. We also expected 88% power to detect group differences in the change between acute and subsequent assessments (Post-RTP, 6-Month) with 40 subjects per group^15^.

### Instrumented Balance Assessments and Measured Outcomes

All procedures were part of a more extensive protocol investigating reactive balance in collegiate athletes^15^. All testing was conducted in an athletic training room setting. Athletes performed the balance task in shoes; if athletes did not wear shoes to testing (e.g., they wore sandals), testing was completed in socks or barefoot.

Participants wore five inertial measurement units (IMUs; Opals v2; APDM Inc., Portland, OR) throughout the balance assessments. Sensors, sampling at 128Hz, were located on top of the metatarsals of the athlete’s left and right feet, on the anterior shank, the lumbar region of the spine (∼L3/L4), and the mid-point of the sternum. The tests were also video recorded with an iPad for reference during data processing.

Reactive balance and compensatory stepping behavior were assessed with the *instrumented-modified Push and Release* (I-mP&R)^18–20^. This clinically viable test is commonly performed as part of the BESTest and mini-BEST for balance assessment in clinical populations^21^. Participants performed four directions (forward, backward, left, and right) in both a single-task condition and a dual-task condition, for a total of eight trials^15,18^. Each participant was randomized to complete the single– or dual-task condition first, and the direction order was kept for all timepoints. During the forward and backward directions, a footplate was placed in between participants’ feet to standardize their foot placement, while during the left and right directions, the participants’ feet were together. During the I-mP&R, the administrator wore an IMU on their right hand to determine the release instant when processing the data^15^. Participants were instructed to lean into the administrator’s hands placed on the participant’s shoulders. The administrator leaned participants until their center of mass was just outside their base of support. This inflection point was then held as participants were instructed to close their eyes, and at a random time (∼2-5s), the administrator rapidly released the participant and participants were allowed to open their eyes. Participants were, thereafter, required to regain their balance and avoid a fall by whatever means necessary. Participants were informed that they could open their eyes after they could feel the administrator release. During the dual-task condition, participants were asked to begin a cognitive task after closing their eyes and were released while completing it. The dual-tasks included serial subtraction by 3’s, reciting the alphabet by every other letter, FAS test, and category recital (animals or fruit). The performance of the participants’ cognitive tasks was not recorded.

IMU recordings were processed through custom algorithms in MATLAB (r2023b; MathWorks, Natick, MA), handling raw accelerations and angular velocities. The primary reactive balance outcome from the I-mP&R^18^ was *time to stability* (in seconds, s), defined as the time from release of support (*t_0_*), to stabilization. The release of support was detected when the administrator’s hand acceleration was >1.05 times gravity. Stabilization was detected when lumbar acceleration was less than 1.07 times gravity and rotational rate was <14°/s after the last step^20^. The secondary outcome was *step latency* (in milliseconds, ms), defined as the time between *t_0_* and the first foot movement, identified when foot acceleration was greater than 1.07 times gravity and the rotational rate was >7°/s^20^. Tertiary exploratory outcomes included *step length* (in meters/meters), defined as the distance of the first step and normalized to the participant’s height; and *time to first contact* (in seconds) defined as the time from *t_0_* and the first step^15^. To maximize reliability, summary metrics of the I-mP&R across all four directions were used instead of direction-specific measures. These summary metrics were the median time to stability, maximum step latency, median time to first contact, and median step length^18^.

### Statistical analysis

Statistical analysis was performed with RStudio (2023.12.0 Build 369; Rstudio Team, 2020, Boston, Ma), following the pre-registered procedure^15^. All analysis were based on all available data. Two sample t-tests with Bonferroni adjustments (5 timepoints; significance level = 0.05 / 5) were used to determine differences between concussed and control subjects at each timepoint.

Considering the non-uniform attendance of participants, we implemented Generalized Estimating Equations (GEEs), which accounts for random missing data^22^, to evaluate the acute and longitudinal effects of concussion on reactive balance. GEEs were defined for time to stability and latency. Fixed effects variables were the assessment timepoint (Baseline, Acute, Pre-RTP, Post-RTP, 6-Month), group (concussion, control) and task (single-task, dual-task), as well as their two-and three-way interactions. Retaining significant interactions at the 0.10 level, the final GEE model included two-way group interactions with assessment (group x assessment timepoint) and task (group x task). All models were also adjusted for covariates of age, sex, height, type of sport (contact or non-contact), and footwear worn during the testing (shoes, barefoot, socks)^23^. Post-hoc pair-wise contrasts compared concussion and control groups at each timepoint when adjusting for covariates.

Following visual inspection of the results, an exploratory post-hoc analysis stratified participants by concussion history to explore potential differences in how lifetime concussion history affected the acute and longitudinal response to a *new* concussion (i.e., the enrollment concussion). In this case, athletes enrolled as concussed subjects without a lifetime history of concussion experienced their first lifetime concussion. Athletes enrolled as concussed subjects with a previous lifetime history of concussion had at least one previous lifetime concussion. GEE models with equivalent fixed effects variables (assessment timepoint, group and task), two-way interactions (group x assessment timepoint and group x task) and covariates (age, sex, height, type of sport, and footwear) were defined for athletes enrolled into the concussion and control groups, stratified by their lifetime history of concussion before enrollment (or the concussion event that lead to enrollment).

To further investigate potential mechanisms underlying differences in our primary outcomes, the relationship between primary and secondary I-mP&R metrics of time to stability and step latency were evaluated using linear regression models and the coefficient of determination, R^2^, for single– and dual-task, in concussed and control groups. A 0.05 significance level was used for all GEE models and exploratory analyses.

## RESULTS

Demographic and characteristics of the involved subjects is reported in Table 1. Overall, 91 athletes were enrolled into this longitudinal arm of the study (47 concussed athletes and 44 healthy teammate controls). Not every subject could be present at all timepoints, with comparable missing data between concussed athletes and matched controls. High rates of lost to follow up at the six months’ timepoint were attributable to Covid-19 pandemic. Groups were overall similar in distribution of sex, age, height, mass, and participation in contact sports, with 78% of the participants being white. At all timepoints, more than half of the athletes enrolled in the concussion group (59.6%) had a history of previous concussions defined as at least one lifetime concussion in addition to their enrollment concussion, while only 47.7% control subjects had history of previous concussions prior to enrollment. Similarly, 48.9% of the concussed athletes had history of lower extremity musculoskeletal injury in the two years before enrollment, while only 31.8% of controls reported a lower extremity musculoskeletal injury within two years before enrollment.

**Table 1.**
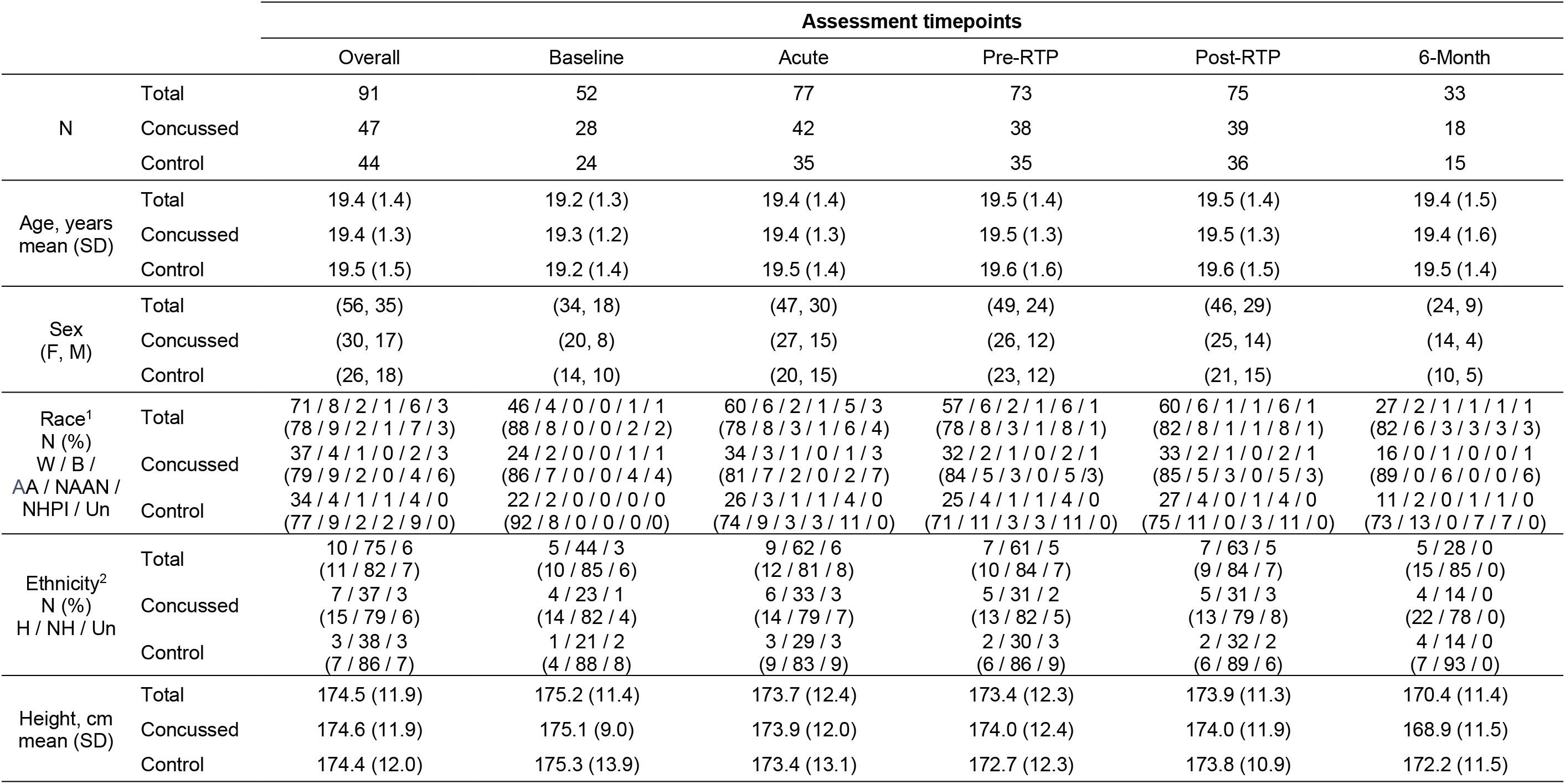
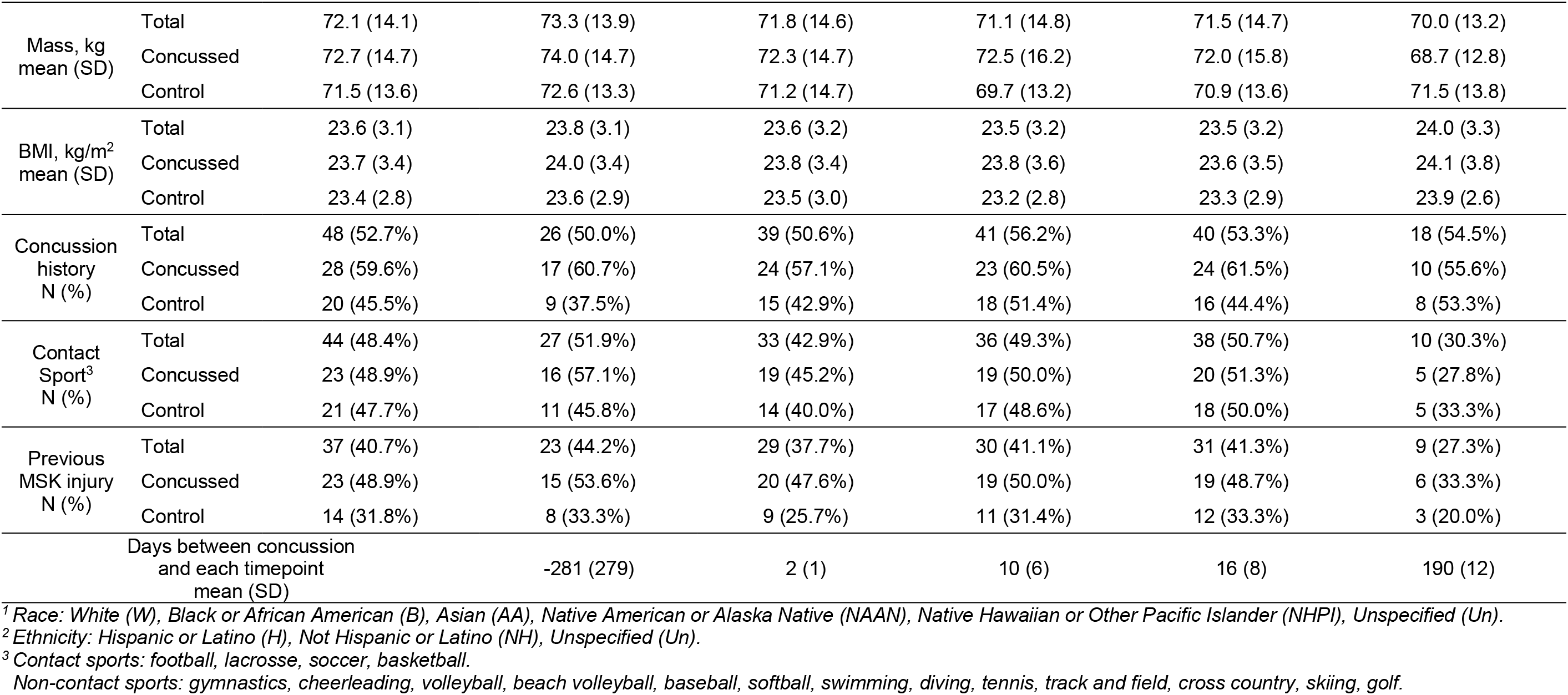
Demographics and characteristics of the population involved (where: N number, s seconds, cm centimeters, kg kilograms, m eters). Mean and standard deviation (SD) are reported for age, weight, body mass index (BMI). Number and percentages are reported for istory of concussion, contact sport and previous musculoskeletal (MSK) injury.

To enhance clarity and streamline the discussion, emphasis is placed on primary and secondary outcomes of time to stability and step latency. Summary statistics of these I-mP&R reactive balance outcomes are reported in Table 2. Supplementary material includes the results for time to first step, and step length.

**Table 2.**
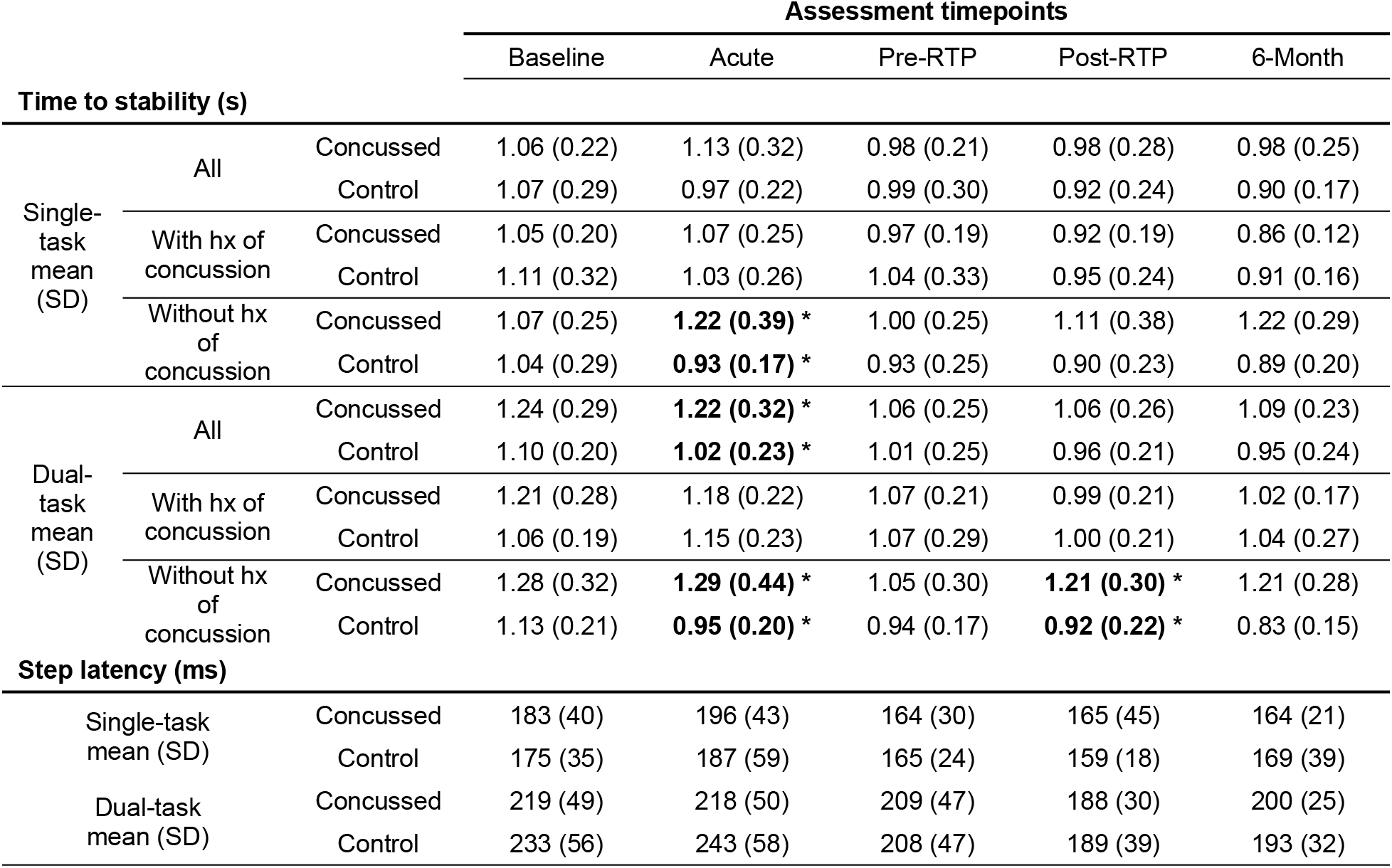
I-mP&R reactive balance outcomes. Mean and standard deviation (SD) of time to stability (s) and step latency (ms) for single-task and dual-task at each timepoint. Significance of Bonferroni adjusted t-test are bold and highlighted with * (α<0.01).

### Time to stability

Bonferroni adjusted cross-sectional t-tests (α = 0.01) revealed athletes with a recent concussion had longer time to stability than healthy controls at the acute timepoint in the dual-task (*p* = 0.007), but not single-task (*p* = 0.027) conditions (Figure 1; Figure 2). There were no differences between groups for any other timepoint, including baseline.

**Figure 1.**
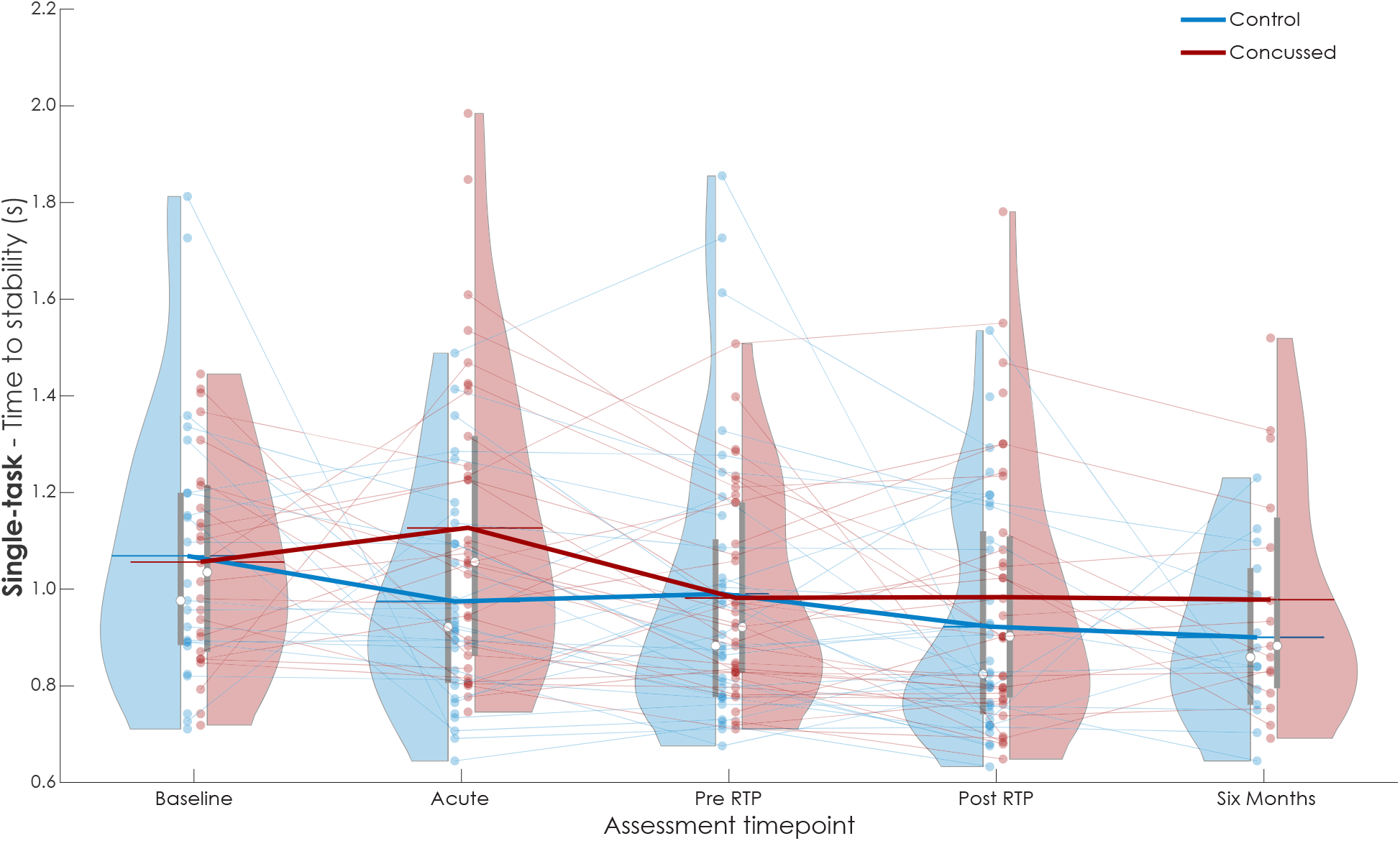
Time to stability (s) for single-task I-mP&R at each assessment timepoint for concussed (red) and control (blue) subjects. Violin plot reports data distribution, mean (white circle) and interquartile range (gray box). Results of each subject relate with semi-transparent lines; thick lines link the averages of each distribution.

**Figure 2.**
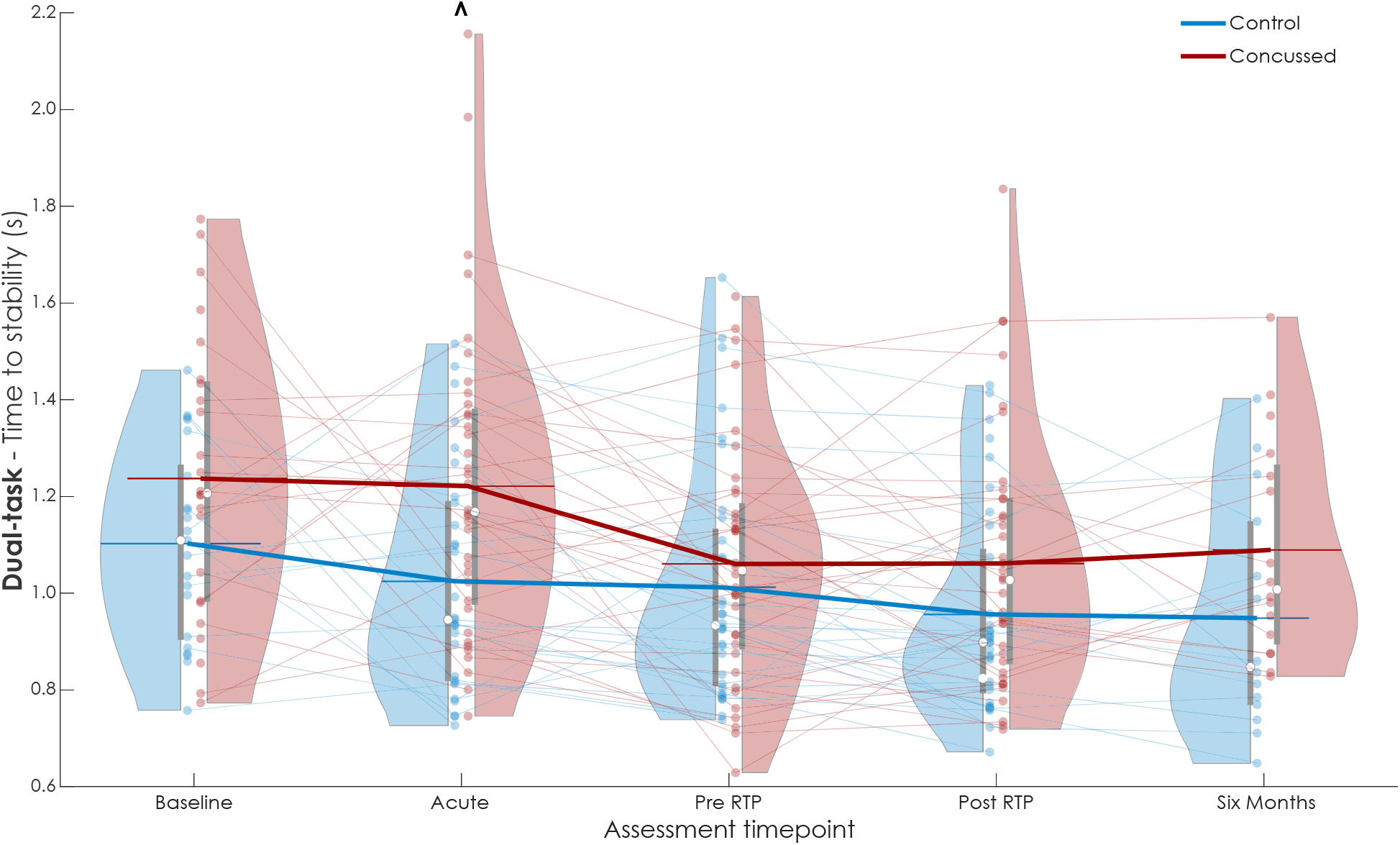
Time to stability (s) for dual-task I-mP&R at each assessment timepoint for concussed (red) and control (blue) subjects. Violin plot reports data distribution, mean (white circle) and interquartile range (gray box). Results of each subject relate with semi-transparent lines; thick lines link the averages of each distribution. ^ highlights significance of the Bonferroni adjusted t-test.

When adjusting for covariates using the GEE framework (Table 3), a main effect of task (*p* = 0.012) and a group*task interaction (*p* = 0.013) indicated both groups exhibited longer time to stability (i.e., worse reactive balance) in the dual-task condition, with the concussion group exhibiting greater increases in time to stability across tasks. Pair-wise contrasts at each time indicated significant group differences at the Acute timepoint for dual-task time to stability (*p*= 0.004), where the concussion group exhibited longer time to stability compared to the control group. There were no other significant differences when adjusting for covariates for single-task (Baseline *p* = 0.940; Acute *p* = 0.051; Pre-RTP *p* = 0.643; Post-RTP *p* = 0.493; 6-Month *p* = 0.401) or dual-task (Baseline *p* = 0.390; Pre-RTP *p* = 0.613; Post-RTP *p* = 0.097; 6-Month *p* = 0.121).

**Table 3.** Generalized estimating equation (GEE) results for time to stability (s) and step latency (ms), reporting for each covariate estimated coefficient β, confidence interval CI, and p value. Reference level for each factor: control for group, Acute for timepoint, single-task for task, female for sex, contact sport for type of sport, shoes for footwear.

|  | Time to stability (s) |  | Step latency (ms) |  |
| --- | --- | --- | --- | --- |
| | $\beta$ (95% CI) | p value | $\beta$ (95% CI) | p value |
| (Intercept) | 0.503 (-0.383, 1.389) | 0.266 | 158 (29, 2867) | 0.016 |
| Time |  |  |  |  |
| Baseline | 0.097 (0.012, 0.182) | 0.026 | -11 (-29, 7) | 0.219 |
| Acute | Ref |  | Ref |  |
| Pre-RTP | -0.013 (-0.082, 0.055) | 0.702 | -28 (-44, -12) | <0.001 |
| Post-RTP | -0.068 (-0.141, 0.006) | 0.070 | -42 (-59, -26) | <0.001 |
| 6-Month | -0.099 (-0.189, -0.009) | 0.030 | -32 (-52, -12) | 0.002 |
| Group |  |  |  |  |
| Control | Ref |  | Ref |  |
| Concussed | 0.110 (-0.001, 0.222) | 0.051 | -3 (-23, 17) | 0.773 |
| Task |  |  |  |  |
| Single-task | Ref |  | Ref |  |
| Dual-task | 0.038 (0.008, 0.068) | 0.012 | 42 (31, 53) | <0.001 |
| Sex |  |  |  |  |
| Female | Ref |  | Ref |  |
| Male | -0.135 (-0.229, -0.041) | 0.005 | 5 (-11, 21) | 0.554 |
| Age | 0.020 (-0.003, 0.043) | 0.086 | 2 (-1, 5) | 0.142 |
| Height | 0.001 (-0.003, 0.004) | 0.670 | -0 (-1, 1) | 0.951 |
| Sport type |  |  |  |  |
| Contact sport | Ref |  | Ref |  |
| Non-contact sport | 0.025 (-0.075, 0.125) | 0.618 | -9 (-22, 3) | 0.145 |
| Footwear |  |  |  |  |
| Shoes | Ref |  | Ref |  |
| Socks | 0.060 (-0.098, 0.217) | 0.459 | -8 (-21, 6) | 0.275 |
| Barefoot | 0.001 (-0.066, 0.068) | 0.972 | -1 (-12, 11) | 0.918 |
| Time * Group |  |  |  |  |
| Baseline * Concussed | -0.115 (-0.276, 0.046) | 0.161 | 6 (-20, 32) | 0.633 |
| Acute * Concussed | Ref |  | Ref |  |
| Pre-RTP * Concussed | -0.138 (-0.245, -0.030) | 0.012 | 8 (-16, 32) | 0.512 |
| Post-RTP * Concussed | -0.064 (-0.172, 0.044) | 0.248 | 12 (-13, 37) | 0.352 |
| 6-Month * Concussed | -0.045 (-0.189, 0.100) | 0.543 | 7 (-21, 36) | 0.619 |
| Task * Group |  |  |  |  |
| Single-task * Concussed | Ref |  | Ref |  |
| Dual-task * Concussed | 0.055 (0.012, 0.099) | 0.013 | -10 (-23, 4) | 0.160 |

When stratified by history of concussion, Bonferroni-corrected t-tests indicated that athletes with their first lifetime concussion (i.e., athletes in the concussion group without a prior history of concussion before the enrollment concussion) exhibited longer times to stability at the Acute timepoint for both single-task (*p* = 0.010) and dual-task (*p* = 0.006) conditions compared to athletes with no lifetime history of concussion enrolled in the study as controls (Figure 3; Figure 4). These dual-task differences between concussed athletes and healthy controls, both without a prior history of concussion before the enrollment event, were also evident during the Post-RTP (*p* = 0.006) and 6-Month (*p* = 0.017) timepoints. In contrast, no differences were observed between concussed and control (all *p* > 0.177) athletes who reported a history of concussion prior to the enrollment event (i.e., all had at least one lifetime concussion before enrollment)

**Figure 3.**
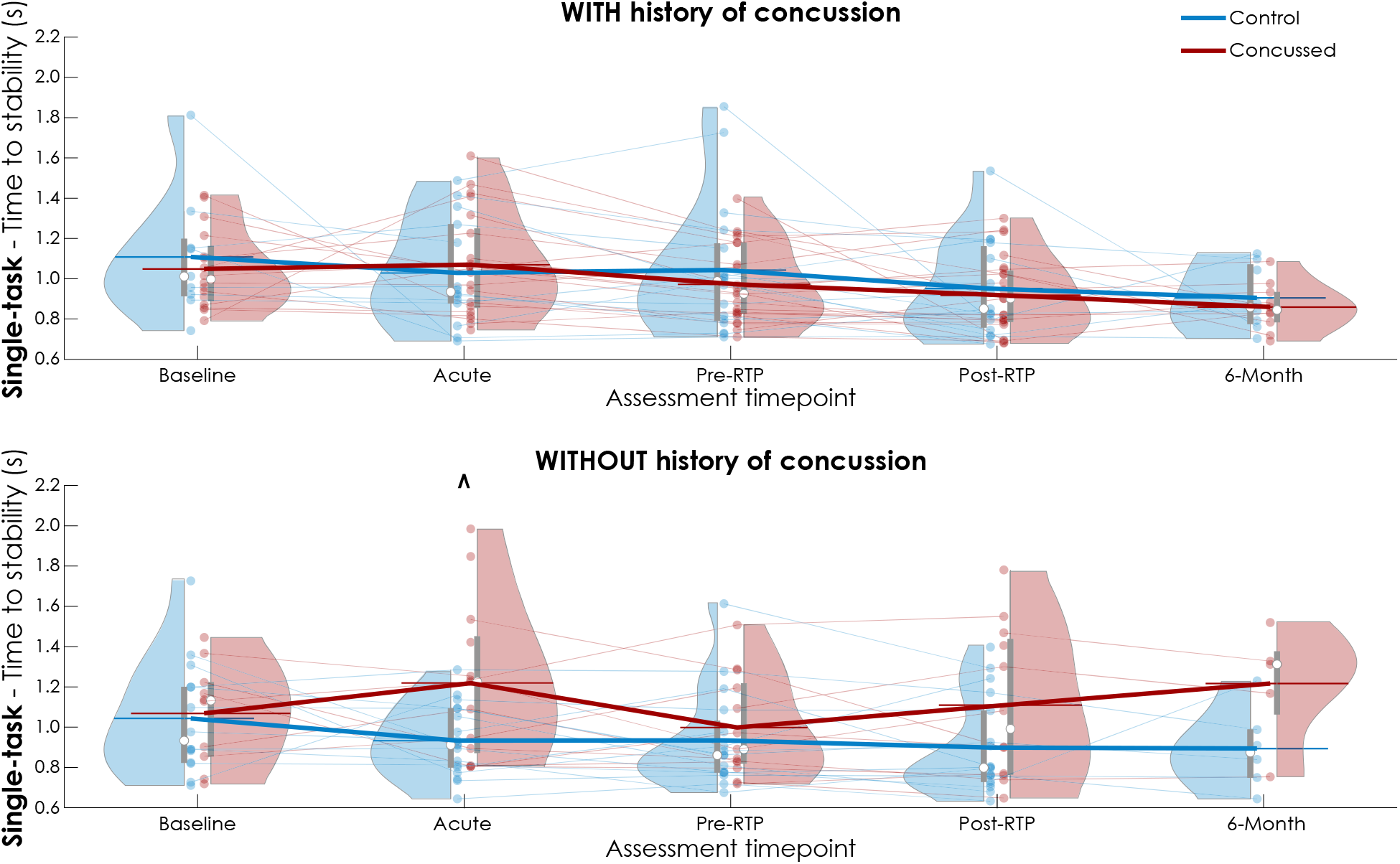
Time to stability (s) for single-task I-mP&R stratified by history of concussion prior to enrollment, at each assessment timepoint for concussed (red) and control (blue) subjects. Violin plot reports data distribution, mean (white circle) and interquartile range (gray box). Results of each subject relate with semi-transparent lines; thick lines link the averages of each distribution. ^ highlights significance of the Bonferroni adjusted t-test.

**Figure 4.**
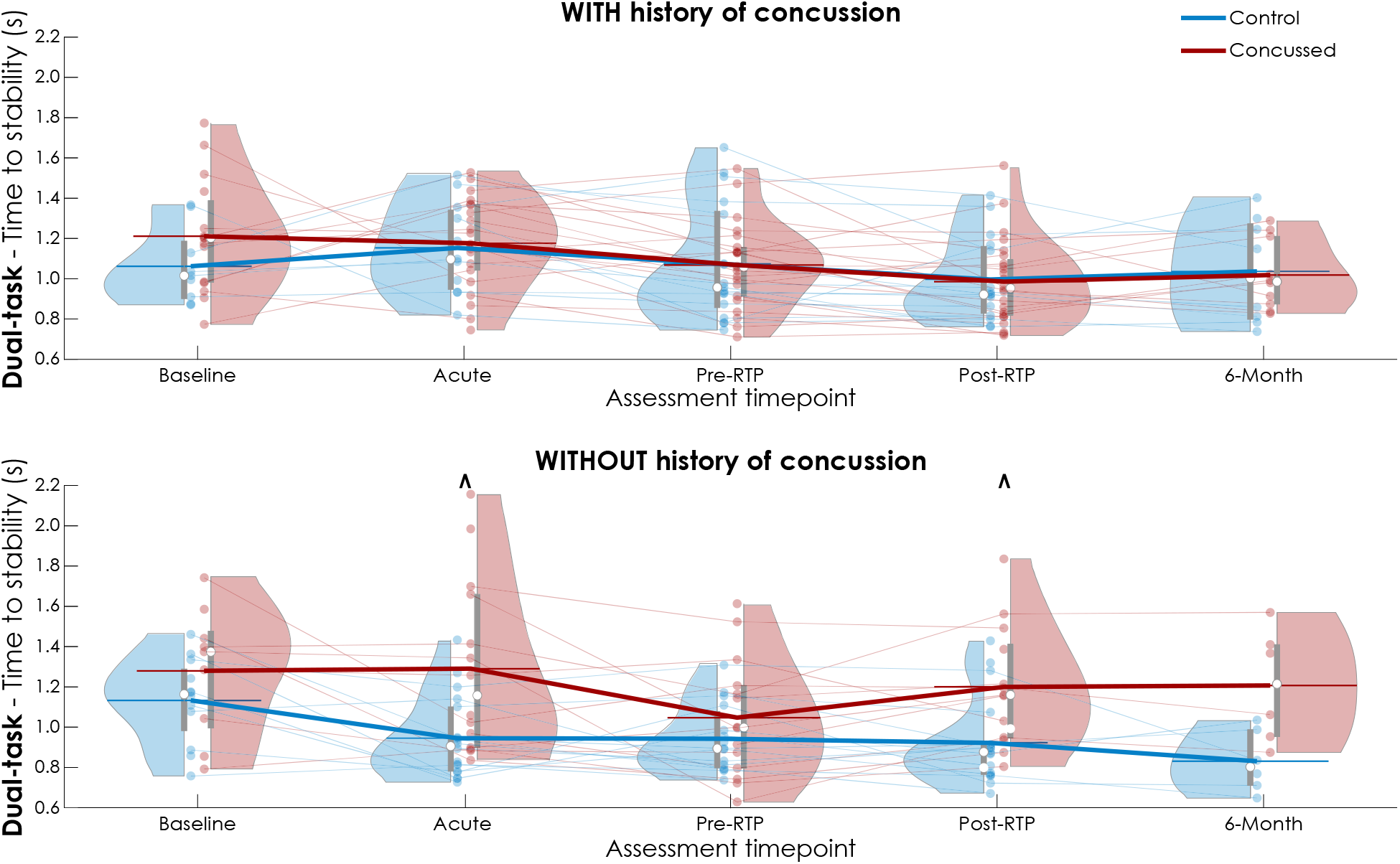
Time to stability (s) for dual-task I-mP&R stratified by history of concussion prior to enrollment, at each assessment timepoint for concussed (red) and control (blue) subjects. Violin plot reports data distribution, mean (white circle) and interquartile range (gray box). Results of each subject relate with semi-transparent lines; thick lines link the averages of each distribution. ^ highlights significance of the Bonferroni adjusted t-test.

Similar results were observed when adjusting for covariates in a GEE framework (Table 4). Pair-wise contrasts revealed concussed athletes without a prior concussion history (i.e., first lifetime concussion) exhibited longer single-task time to stability at the Post-RTP timepoint (*p* = 0.035), and longer dual-task times to stability at Acute (*p* = 0.037) and Post-RTP (*p* = 0.004) timepoints compared to athletes with no lifetime concussion. Notable, but non-significant effects were also observed at 6-Months for both single-task (*p* = 0.073) and dual-task (*p* = 0.054). Similar to the unadjusted t-tests, no differences were observed between the concussion and control athletes with a lifetime history of concussion before enrollment (all *p* > 0.414).

**Table 4.** Generalized estimating equation (GEE) of time to stability (s), stratified by history of concussion, reporting for each covariate estimated coefficient β, confidence interval CI, and p value. Reference level for each factor: control for group, Acute for timepoint, single-task for task, female for sex, contact sport for type of sport, shoes for footwear.

|  | Time to stability (s) |  |  |  |
| --- | --- | --- | --- | --- |
|  | With history of concussion<br>(n = 48) |  | Without history of concussion<br>(n = 43) |  |
| | $\beta$ (95% CI) | p value | $\beta$ (95% CI) | p value |
| (Intercept) | 1.662 (0.229, 3.095) | 0.023 | -0.262 (-1.598, 1.074) | 0.701 |
| Time |  |  |  |  |
| Baseline | 0.035 (-0.128, 0.197) | 0.677 | 0.143 (0.032, 0.254) | 0.011 |
| Acute | Ref |  | Ref |  |
| Pre-RTP | -0.054 (-0.123, 0.016) | 0.133 | -0.027 (-0.120, 0.066) | 0.575 |
| Post-RTP | -0.130 (-0.257, -0.002) | 0.046 | -0.045 (-0.119, 0.029) | 0.231 |
| 6-Month | -0.152 (-0.228, -0.077) | <0.001 | -0.072 (-0.168, 0.024) | 0.143 |
| Group |  |  |  |  |
| Control | Ref |  | Ref |  |
| Concussed | 0.014 (-0.127, 0.155) | 0.846 | 0.168 (-0.039, 0.374) | 0.112 |
| Task |  |  |  |  |
| Single-task | Ref |  | Ref |  |
| Dual-task | 0.057 (0.007, 0.108) | 0.026 | 0.025 (-0.008, 0.057) | 0.138 |
| Sex |  |  |  |  |
| Female | Ref |  | Ref |  |
| Male | 0.018 (-0.094, 0.131) | 0.747 | -0.306 (-0.427, -0.184) | <0.001 |
| Age | 0.006 (-0.026, 0.037) | 0.727 | 0.027 (-0.006, 0.060) | 0.110 |
| Height | -0.004 (-0.010, 0.001) | 0.137 | 0.005 (-0.003, 0.012) | 0.208 |
| Sport type |  |  |  |  |
| Contact sport | Ref |  | Ref |  |
| Non-contact sport | 0.021 (-0.088, 0.129) | 0.712 | -0.038 (-0.173, 0.097) | 0.581 |
| Footwear |  |  |  |  |
| Shoes | Ref |  | Ref |  |
| Socks | 0.117 (-0.052, 0.286) | 0.176 | -0.079 (-0.237, 0.078) | 0.322 |
| Barefoot | -0.030 (-0.114, 0.053) | 0.480 | 0.011 (-0.070, 0.092) | 0.798 |
| Time * Group |  |  |  |  |
| Baseline * Concussed | -0.026 (-0.230, 0.179) | 0.806 | -0.154 (-0.394, 0.085) | 0.206 |
| Acute * Concussed | Ref |  | Ref |  |
| Pre-RTP * Concussed | -0.063 (-0.169, 0.043) | 0.245 | -0.142 (-0.327, 0.042) | 0.131 |
| Post-RTP * Concussed | -0.045 (-0.184, 0.093) | 0.519 | 0.034 (-0.121, 0.190) | 0.664 |
| 6-Month * Concussed | -0.008 (-0.125, 0.110) | 0.897 | 0.057 (-0.218, 0.333) | 0.684 |
| Task * Group |  |  |  |  |
| Single-task * Concussed | Ref |  | Ref |  |
| Dual-task * Concussed | 0.049 (-0.009, 0.108) | 0.098 | 0.042 (-0.021, 0.106) | 0.193 |

### Step latency

No statistically significant group differences in latency were observed using Bonferroni adjusted t-tests (all *p* > 0.067) or when adjusting for covariates in the GEE framework (Table 3) for single-task (Supplement Figure S1) or dual-task conditions (Supplement Figure S2), (*p* > 0.128 at all timepoints). Within the GEE framework, a main effect of task was observed (*p* < 0.001), where latency was longer in dual-task conditions compared to single-task conditions, but there was no significant group * task interaction (*p* = 0.160).

### Time to stability vs Step latency

There was no significant relationship between time to stability and step latency in any group or condition at the Acute timepoint (Figure 5). Linear relationships ranged from R^2^ = 0.005 for controls during dual-task conditions to R^2^ = 0.091 for controls during single-task.

**Figure 5.**
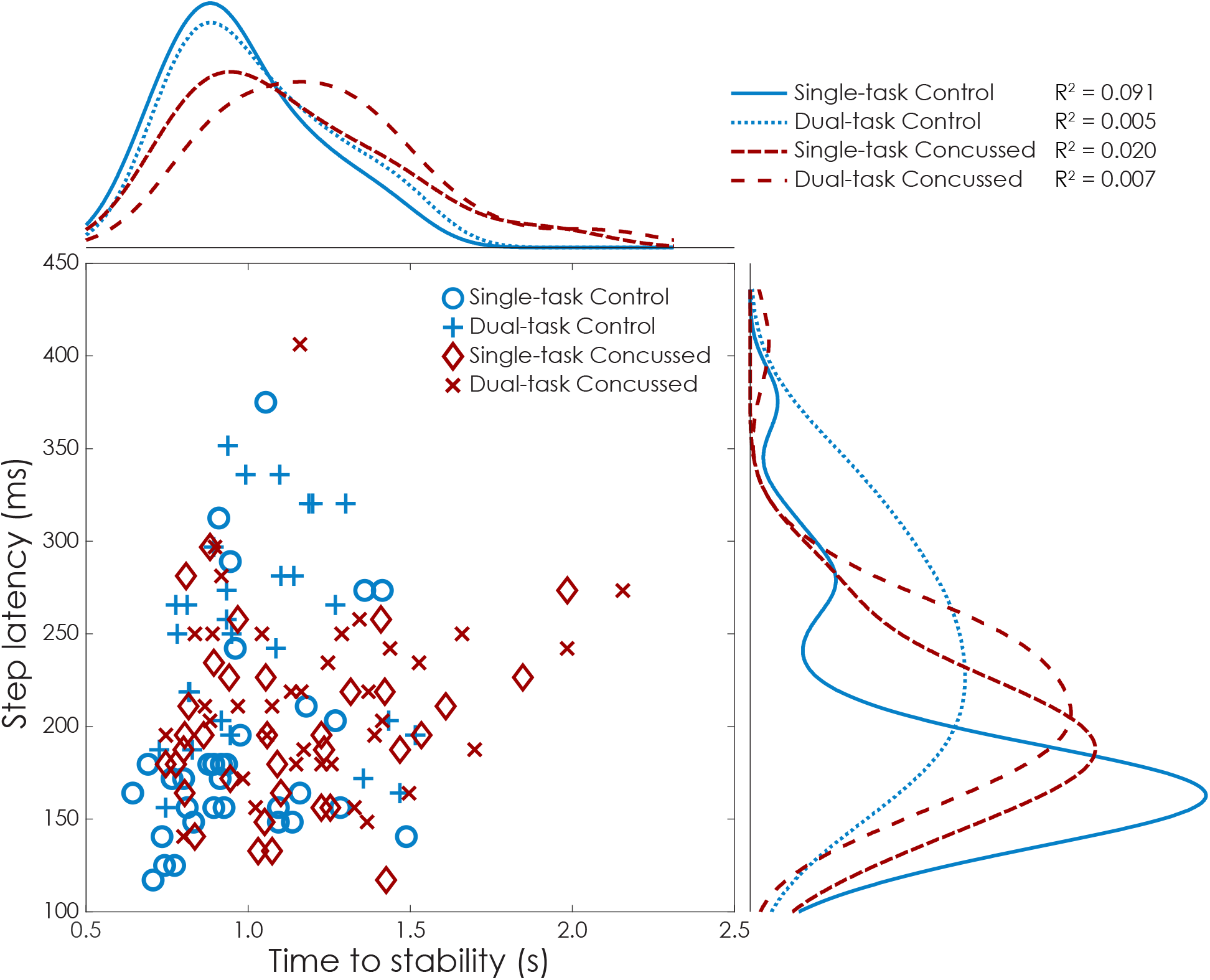
Scatter histogram of the relationship between time to stability (s) and step latency (ms) at acute timepoint. For single– and dual-task, concussed (red) and control (blue) populations are reported distributions curves and R^2^ regression coefficients.

## DISCUSSION

This study explored the longitudinal effects of concussion on reactive balance in college athletes. Acutely after concussion (i.e., < 72 hours), athletes exhibited impaired reactive balance, characterized by longer times to stability in dual-task conditions. We observed no group differences in time to stability at baseline, providing strong evidence that the slower time to stabilities at post-concussion timepoints were associated with the concussion injury. When further adjusting for demographic (age, sex, height, sport type) and test-specific covariates (footwear), we still observed acute impairments in dual-task reactive balance. These impairments in reactive balance were transient; athletes did not exhibit overall group differences at longitudinal assessments outside the Acute timepoint. However, we observed differential effects of concussion on reactive balance based on one’s lifetime history of concussion; athletes who suffered their first lifetime concussion exhibited acute and longitudinal impairments in both single– and dual-task reactive balance compared to their peers with no lifetime concussion.

Dual-task performance differences between concussed and control athletes align with previous studies emphasizing the importance of attentional resources in managing cognitive and motor demands^24^. The link between cognitive processing and postural stability is challenged during dual-task scenarios^12^, where simultaneous engagement strains attentional capacity, affecting performance^25^. Dual-task paradigms have also proven to be more effective in challenging athletes, possibly explaining the differences that were observed exclusively in dual-task conditions rather than single-task situations. Additionally, similar tests on non-athletes might yield different results due to the specialized training, skills and familiarity with reactions tasks of athletes.

The differences between groups at the acute timepoint were not solely due to a regression, relative to baseline, in reactive balance performance in the concussion group – deficits appeared to be driven by the concussion group getting slower *and* the control group getting faster relative to baseline. Such improvements in the control group across time are likely indicative of learning and adaptation effects commonly observed in assessments of reactive balance^26,27^. Considering the large time from baseline to acute timepoints (mean 281 days), it is also possible such improvements in reactive balance in health controls reflect overall improvements in motor function and postural control – including reactive balance – through the balance, strength, and conditioning training integrated into collegiate programs. In contrast, athletes with concussion did not exhibit improved performance relative to baseline until they were asymptomatic (Pre-RTP timepoint). These findings suggest that concussion have repercussions on learning effects seen in the control group^28,29^.

While the mechanisms underlying the acute post-concussion deficits in reactive balance remain unclear, it is unlikely that reaction time is an underlying cause. Slower reaction times are well-documented after concussion using both computerized neurocognitive testing and clinical drop-stick tests^30^. Yet, these traditional measures of reaction time are not associated with postural reaction times^13,23^ (e.g., step latency). We did not observe differences in step latency across groups at any timepoint, and there were negligible associations between step latency and time to stability at the acute timepoint. Further, a notable but non-significant interaction, was evident where athletes with acute concussion exhibited *slower* time to stability, yet *faster* step latencies, in the dual-task condition compared to their healthy teammates. Combined, such results across time and (lack of) associations with step latency suggest post-concussion reactive balance impairments in time to stability may be associated with altered trans-cortical loops, rather than short– or medium-latency responses. Altered long-latency trans-cortical responses may be influenced by an inability to appropriately prime a stepping response before the release^31^ or an inappropriate motor responses, driven by altered sensory integration and state estimation, after a loss of balance.^14,32^ However, such conclusions remain speculative and further research is needed to uncover the mechanisms underlying concussion-related reactive balance impairments.

Stratifying time to stability based on concussion history indicated that the acute reactive balance impairments are influenced by one’s lifetime history of concussion. Significant group differences were observed in athletes without a history of concussion acutely and, for dual-task, at Post-RTP and 6-Month timepoints. While comparisons across the four groups were omitted due to insufficient statistical power and misalignment with the study’s scope, descriptive comparisons revealed that athletes in the control group, who did not experience any lifetime concussions, exhibited the fastest time to stability amongst all groups. Conversely, concussed with a history of concussion did not differ from control athletes with a history of concussion, and both groups exhibited worse performance during dual-task conditions, indicating that deficits from previous concussions may persist over time^33^. Declines in dual-task performance are linked to an increased risk of musculoskeletal injury^10^, particularly in individuals with worsened abilities relative to baseline^34^ or those exhibiting regression during the return-to-play protocol^35^.

The primary limitation of the presented study is the non-uniform attendance at each assessment, potentially introducing bias from fluctuating sample sizes and affecting the generalizability of our findings. Nonetheless, the GEE model is advantageous for handling missing data, assuming completely at random missing data mechanism, allowing for a robust analysis of longitudinal observations^22^. Additionally, noisy assessment environment and inter-administrator variability may have affected the results, although they reflect clinical settings of concussion assessments and a previous investigation reported moderate inter-administrator reliability and high validity of the I-mP&R^18^. The comprehensive understanding of the neurophysiological mechanisms influencing reactive balance was also compromised by the absence of muscle activity data. Addressing these constraints, future research should enhance the robustness of longitudinal assessments by directly investigating the effects of concussion on the ability to prime (i.e., prepare) a motor response and the ability to execute said motor response. The identified long-term deficits and the importance of history of concussion in reactive balance, particularly in dual-task scenarios, emphasize the need to expand traditional evaluations incorporating reactive balance assessments into concussion protocols.

## CONCLUSION

Athletes demonstrated acute, but transient, reactive balance impairments in dual-task conditions. While the mechanisms of these impairments remain unclear, such impairments are unlikely to be attributed to slower reaction times. Additionally, the effects of concussion on reactive balance were influenced by concussion history; athletes experiencing their first lifetime concussion exhibited both acute and longitudinal impairments in single– and dual-task reactive balance compared to control athletes with no lifetime concussion. Such persistent reactive balance impairments may be associated with the increased risk of musculoskeletal injury risk post-concussion. Future work should further explore the mechanistic origins and clinical significance (e.g., injury risk) of reactive balance impairments after concussion.

## Data Availability

All data produced in the present study are available upon reasonable request to the authors

## ACKNOWLEDGEMENTS

The authors would like to acknowledge the contributions of Benjamin Cassidy, Ryan Pelo, Nicholas Kreter, Sarah Hill, Brody Roemmich, Christina Geisler, Jun Son, Cameron Jensen, Corinne Mayfield, and the University of Utah Athletic Training Staff.

## REFERENCES

1. Harmon KG, Clugston JR, Dec K, et al. American Medical Society for Sports Medicine Position Statement on Concussion in Sport. Clin J Sport Med. Mar 2019;29(2):87–100. doi:10.1097/JSM.0000000000000720

2. Horak FB. Postural orientation and equilibrium: what do we need to know about neural control of balance to prevent falls? Age Ageing. Sep 2006;35 Suppl 2:ii7–ii11. doi:10.1093/ageing/afl077

3. Horak FB. Postural orientation and equilibrium: what do we need to know about neural control of balance to prevent falls? Age Ageing. Sep 2006;35:7–11. doi:10.1093/ageing/afl077

4. Horak FB, Wrisley DM, Frank J. The Balance Evaluation Systems Test (BESTest) to Differentiate Balance Deficits. Phys Ther. May 2009;89(5):484–498. doi:10.2522/ptj.20080071

5. Morris A, Casucci T, McFarland MM, et al. Reactive Balance Responses After Mild Traumatic Brain Injury: A Scoping Review. J Head Trauma Rehab. Sep-Oct 2022;37(5):311–317. doi:10.1097/Htr.0000000000000761

6. Condon C, Cremin K. Static balance norms in children. Physiother Res Int. Mar 2014;19(1):1–7. doi:10.1002/pri.1549

7. Davlin CD. Dynamic balance in high level athletes. Percept Mot Skills. Jun 2004;98(3 Pt 2):1171–6. doi:10.2466/pms.98.3c.1171-1176

8. Pollock AS, Durward BR, Rowe PJ, Paul JP. What is balance? Clin Rehabil. Aug 2000;14(4):402–6. doi:10.1191/0269215500cr342oa

9. Kim Y, Vakula MN, Bolton DAE, et al. Which Exercise Interventions Can Most Effectively Improve Reactive Balance in Older Adults? A Systematic Review and Network Meta-Analysis. Front Aging Neurosci. Jan 18 2022;13 doi: ARTN76482610.3389/fnagi.2021.764826

10. Morris A, Fino NF, Pelo R, et al. Reactive postural responses predict risk for acute musculoskeletal injury in collegiate athletes. J Sci Med Sport. Feb 2023;26(2):114–119. doi: 10.1016/j.jsams.2023.01.003

11. Takakusaki K. Functional Neuroanatomy for Posture and Gait Control. J Mov Disord. Jan 2017;10(1):1–17. doi:10.14802/jmd.16062

12. Maki BE, McIlroy WE. Cognitive demands and cortical control of human balance-recovery reactions. J Neural Transm (Vienna). 2007;114(10):1279–96. doi:10.1007/s00702-007-0764-y

13. Morris A, Petersell TL, Pelo R, et al. Use of Reactive Balance Assessments With Clinical Baseline Concussion Assessments in Collegiate Athletes. J Athl Train. Jan 1 2024;59(1):39–48. doi:10.4085/1062-6050-0231.22

14. Pan T, Liao K, Roenigk K, Daly JJ, Walker MF. Static and dynamic postural stability in veterans with combat-related mild traumatic brain injury. Gait Posture. Oct 2015;42(4):550–557. doi:10.1016/j.gaitpost.2015.08.012

15. Morris A, Cassidy B, Pelo R, et al. Reactive Postural Responses After Mild Traumatic Brain Injury and Their Association With Musculoskeletal Injury Risk in Collegiate Athletes: A Study Protocol. Front Sports Act Living. 2020;2:574848. doi:10.3389/fspor.2020.574848

16. Bohr AD, Aukerman DF, Harmon KG, et al. Pac-12 CARE-Affiliated Program: structure, methods and initial results. BMJ Open Sport Exerc Med. 2021;7(2):e001055. doi:10.1136/bmjsem-2021-001055

17. McCrory P, Meeuwisse W, Dvorak J, et al. Consensus statement on concussion in sport-the 5(th) international conference on concussion in sport held in Berlin, October 2016. Br J Sports Med. Jun 2017;51(11):838–847. doi:10.1136/bjsports-2017-097699

18. Morris A, Fino NF, Pelo R, et al. Interadministrator Reliability of a Modified Instrumented Push and Release Test of Reactive Balance. J Sport Rehabil. May 1 2022;31(4):517–523. doi:10.1123/jsr.2021-0229

19. Smith BA, Carlson-Kuhta P, Horak FB. Consistency in Administration and Response for the Backward Push and Release Test: A Clinical Assessment of Postural Responses. Physiother Res Int. Mar 2016;21(1):36–46. doi:10.1002/pri.1615

20. El-Gohary M, Peterson D, Gera G, Horak FB, Huisinga JM. Validity of the Instrumented Push and Release Test to Quantify Postural Responses in Persons With Multiple Sclerosis. Arch Phys Med Rehabil. Jul 2017;98(7):1325–1331. doi:10.1016/j.apmr.2017.01.030

21. Franchignoni F, Horak F, Godi M, Nardone A, Giordano A. Using psychometric techniques to improve the Balance Evaluation Systems Test: the mini-BESTest. J Rehabil Med. Apr 2010;42(4):323–31. doi:10.2340/16501977-0537

22. Liang KY, Zeger SL. Longitudinal Data-Analysis Using Generalized Linear-Models. Biometrika. Apr 1986;73(1):13–22. doi: DOI 10.1093/biomet/73.1.13

23. Petersell TL, Quammen DL, Crofts R, et al. Instrumented Static and Reactive Balance in Collegiate Athletes: Normative Values and Minimal Detectable Change. J Athl Train. Nov 28 2023; doi:10.4085/1062-6050-0403.23

24. Yogev-Seligmann G, Hausdorff JM, Giladi N. The role of executive function and attention in gait. Mov Disord. Feb 15 2008;23(3):329–42; quiz 472. doi:10.1002/mds.21720

25. Posner MI, Boies SJ. Components of Attention. Psychol Rev. 1971;78(5):391-&. doi: DOI 10.1037/h0031333

26. Welch TDJ, Ting LH. Mechanisms of Motor Adaptation in Reactive Balance Control. Plos One. May 8 2014;9(5) doi: ARTNe9644010.1371/journal.pone.0096440

27. Horak FB, Nashner LM. Central programming of postural movements: adaptation to altered support-surface configurations. J Neurophysiol. Jun 1986;55(6):1369–81. doi:10.1152/jn.1986.55.6.1369

28. Bourassa ME, Dumel G, Charlebois-Plante C, Gagnon JF, De Beaumont L. Persistent implicit motor learning alterations following a mild traumatic brain injury sustained during late adulthood. J Clin Exp Neuropsyc. Jan 2 2021;43(1):105–115. doi:10.1080/13803395.2021.1879735

29. Cantarero G, Choynowski J, St Pierre M, et al. Repeated Concussions Impair Behavioral and Neurophysiological Changes in the Motor Learning System. Neurorehab Neural Re. Sep 2020;34(9):804–813. doi:Artn 154596832094357810.1177/1545968320943578

30. Lempke LB, Howell DR, Eckner JT, Lynall RC. Examination of Reaction Time Deficits Following Concussion: A Systematic Review and Meta-analysis. Sports Med. Jul 2020;50(7):1341–1359. doi:10.1007/s40279-020-01281-0

31. Kreter N, Rogers CL, Fino PC. Anticipatory and reactive responses to underfoot perturbations during gait in healthy adults and individuals with a recent mild traumatic brain injury. *Clin Biomech (Bristol*, Avon). Dec 2021;90:105496. doi:10.1016/j.clinbiomech.2021.105496

32. Campbell KR, King LA, Parrington L, Fino PC, Antonellis P, Peterka RJ. Central sensorimotor integration assessment reveals deficits in standing balance control in people with chronic mild traumatic brain injury. Front Neurol. 2022;13:897454. doi:10.3389/fneur.2022.897454

33. Howell DR, Beasley M, Vopat L, Meehan WP, 3rd. The Effect of Prior Concussion History on Dual-Task Gait following a Concussion. J Neurotrauma. Feb 15 2017;34(4):838–844. doi:10.1089/neu.2016.4609

34. Oldham JR, Howell DR, Knight CA, Crenshaw JR, Buckley TA. Gait Performance Is Associated with Subsequent Lower Extremity Injury following Concussion. Med Sci Sport Exer. Nov 2020;52(11):2279–2285. doi:10.1249/Mss.0000000000002385

35. Howell DR, Buckley TA, Lynall RC, Meehan WP. Worsening Dual-Task Gait Costs after Concussion and their Association with Subsequent Sport-Related Injury. J Neurotraum. Jul 2018;35(14):1630–1636. doi:10.1089/neu.2017.5570

