## Supplemental Figure and Table for "Acute and Longitudinal Effects of Sport-related Concussion on Reactive Balance"

**SUPPLEMENTARY MATERIAL**

*Figure S1. Step latency (ms) for single-task I-mP&R at each assessment timepoint for concussed (red) and control (blue) subjects. Violin plot reports data distribution, mean (white circle) and interquartile range (gray box). Results of each subject relate with semi-transparent lines; thick lines link the averages of each distribution.*

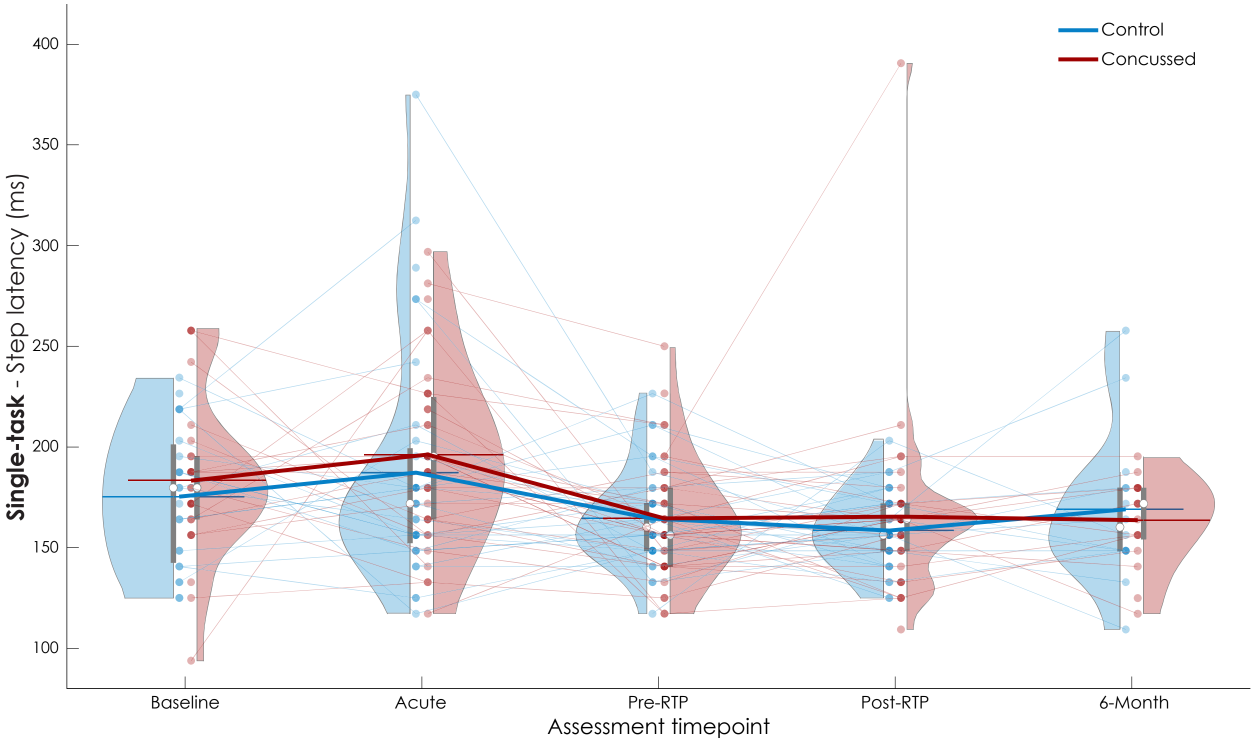

*Figure S2. Step latency (ms) for dual-task I-mP&R at each assessment timepoint for concussed (red) and control (blue) subjects. Violin plot reports data distribution, mean (white circle) and interquartile range (gray box). Results of each subject relate with semi-transparent lines; thick lines link the averages of each distribution.*

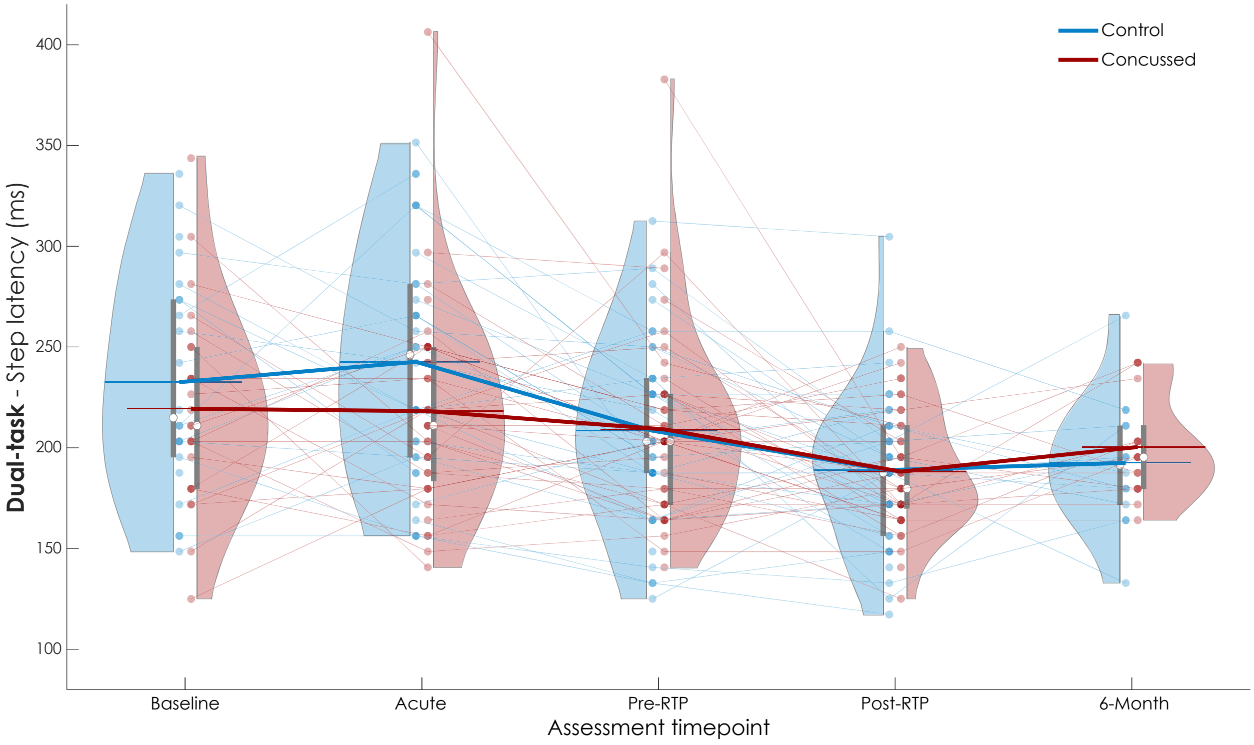

*Table S2. I-mP&R reactive balance outcomes. Mean and standard deviation (SD) of step length, normalized by subjects' height, and time to first contact for single-task and dual-task at each timepoint.*

| | | Assessment timepoints | | | | |
|---|---|---|---|---|---|---|
| | | Baseline | Acute | Pre-RTP | Post-RTP | Six-Months |
| **Step length (m/m)** | | | | | | |
| Single-task mean (SD) | Concussed | 0.28 (0.05) | 0.28 (0.04) | 0.28 (0.04) | 0.28 (0.04) | 0.28 (0.04) |
| | Control | 0.27 (0.04) | 0.29 (0.04) | 0.28 (0.04) | 0.29 (0.03) | 0.29 (0.04) |
| Dual-task mean (SD) | Concussed | 0.27 (0.05) | 0.29 (0.06) | 0.28 (0.05) | 0.28 (0.04) | 0.29 (0.04) |
| | Control | 0.27 (0.04) | 0.29 (0.04) | 0.28 (0.04) | 0.28 (0.04) | 0.28 (0.04) |
| **Time to first foot contact (s)** | | | | | | |
| Single-task mean (SD) | Concussed | 0.44 (0.07) | 0.44 (0.07) | 0.41 (0.06) | 0.41 (0.06) | 0.42 (0.05) |
| | Control | 0.43 (0.06) | 0.43 (0.06) | 0.42 (0.04) | 0.42 (0.06) | 0.41 (0.05) |
| Dual-task mean (SD) | Concussed | 0.50 (0.07) | 0.49 (0.07) | 0.45 (0.06) | 0.44 (0.06) | 0.46 (0.04) |
| | Control | 0.46 (0.07) | 0.48 (0.05) | 0.46 (0.06) | 0.44 (0.06) | 0.45 (0.04) |
